# Theta burst stimulation is not inferior to high frequency repetitive transcranial magnetic stimulation in reducing symptoms of post-traumatic stress disorder in Veterans with depression: a retrospective case series

**DOI:** 10.1101/2022.09.11.22279828

**Authors:** Mohammad Ali Shenasa, Ellerman Em, Canet Phil, Brian Martis, Jyoti Mishra, Dhakshin Ramanathan

## Abstract

**Background:** Two commonly used forms of repetitive transcranial magnetic stimulation (rTMS) were recently shown to be equivalent for the treatment of treatment-resistant depression (TRD): high-frequency stimulation (10 Hz), a protocol that lasts between 19-38 minutes, and intermittent Theta-Burst Stimulation (iTBS), a protocol that can be delivered in just 3 minutes. Intermittent TBS offers significant time advantages to patients and clinics and has thus become a default treatment in many clinics. However, it is unclear whether iTBS treatment offers the same benefits as standard 10 Hz rTMS for comorbid symptoms, such as post-traumatic-stress-disorder (PTSD).

**Methods:** In this retrospective case series, we analyzed treatment outcomes in Veterans from the VA San Diego Healthcare system (VASDHS) who received 10 Hz (n = 47) or iTBS (n = 51) rTMS treatments for TRD between the dates of Feb 2018 to June 2022. We compared outcomes between these two stimulation protocols used between these dates on symptoms of depression (using changes in the patient health questionnaire-9, or PHQ-9) and PTSD (using changes in the PTSD Checklist for DSM-5, or PCL-5). We hypothesized that there would be no differences in treatment outcomes between 10 Hz and iTBS protocols for either depression (confirming prior RCT) or PTSD.

**Results:** We initially found that stimulation groups differed in gender (the iTBS group had 16 females and 35 males, the 10 Hz group had 5 females and 42 males, p<0.003). Thus, to analyze whether there was a difference by stimulation protocol, we first implemented a mixed-effects ANOVA model for PHQ-8 scores with gender and stimulation type as between-group fixed effects and treatment (pre-treatment and post-treatment scores) as the repeated measures factor. We found no significant difference by stimulation protocol for either depression (PHQ-9, (F(1,94)= 0.16, p = 0.69, eta-squared = 0.002) or PTSD symptoms (PCL-5, F (1,94) = 3.46, p = 0.067, eta-squared = 0.036). As differences related to PTSD outcomes were close to significance, we did look at the post-hoc treatment effects by stimulation type for PTSD symptoms. The iTBS group showed a reduction from 41.9 +/- 4.4 to 25.1 +/- 4.9 (a difference of 16.8 points) while the 10Hz group showed a reduction from 43.6 +/- 2.9 down to 35.2 +/- 3.2 (a difference of 8.4 points). Follow-up analyses restricting the sample in various ways did not meaningfully change these results (no follow-up analyses showed that there was a significant difference between stimulation protocols).

**Conclusions:** While limited by small sample size, non-blinded and pseudo-randomized assignment, our data suggests that iTBS is non-inferior to 10Hz stimulation in inducing reductions in PTSD symptoms and depression in military Veterans. Our findings pave the way for further research trials to validate and optimize iTBS for PTSD symptoms.

## Introduction

Repetitive transcranial magnetic stimulation (rTMS) has been cleared by the US Food and Drug Administration (FDA) for treatment resistant depression (TRD) since 2006 (Gaynes et al., 2014; Sehatzadeh et al., 2019). In clinical rTMS treatment protocols for depression, a magnetic coil is placed against the scalp to deliver electromagnetic pulses to the dorsolateral prefrontal cortex (DLFC), a target thought to improve affective regulation and functional connectivity with core nodes of the depression network (Fox et al., 2012). The initial FDA-cleared clinical protocol used a four second train of pulses, delivered at a frequency of 10 Hz (i.e., 40 total pulses), followed by an inter-train “rest” period ranging from 12-26 seconds, for a total of 3000 pulses. These sessions are typically offered 5 days a week for 6 weeks, and are often offered for 20-30 days (George et al., 2010; Lisanby et al., 2009; O’Reardon et al., 2007). Recently, intermittent theta burst stimulation (iTBS, or TBS) has emerged as an FDA-cleared alternative TMS protocol for depression. Intermittent TBS entails delivery of three short pulses (a triplet burst delivered at 50 Hz) repeated every 200 msec (5 Hz, the classical theta rhythm) (Chistyakov et al., 2010; Huang et al., 2005). In a large multicenter trial, iTBS was non-inferior to the standard 10 Hz rTMS treatment in patients with TRD (Blumberger et al., 2018). Intermittent TBS offers benefits primarily due to its shorter treatment duration (3 vs 37.5 minutes) (Blumberger et al., 2018), easing patient burden and improving efficiency and access of this expensive treatment. Further, iTBS allows for accelerated protocols in which multiple treatments can be delivered daily, allowing for more rapid treatment effects (Cole et al., 2020).

Repetitive TMS has also been applied to reduce symptoms of PTSD, a distressful and potentially disabling condition (for comprehensive review see: Petrosino et al., 2021). PTSD is marked by intrusive memories, avoidance of stimuli, negative changes in mood, cognition, arousal, and hyper-reactivity (American Psychiatric Association, 2013). Importantly, most patients with PTSD also suffer from comorbid depression (Flory & Yehuda, 2015), and the interaction between comorbid depression and PTSD has been reported to worsen both conditions (Shalev et al., 1998). There has been conflicting evidence regarding whether comorbid PTSD symptoms/diagnoses affect treatment response of rTMS in the treatment of depression. For example, in one study of patients with depression, response and remission rates after rTMS were similar between those with and without PTSD (Hernandez et al., 2020). However a randomized controlled trial (RCT) reported that remission rates were lower in patients with comorbid PTSD (Yesavage et al., 2018). Despite this conflicting evidence and a lack of FDA clearance for the treatment of PTSD, rTMS has been shown in smaller research studies to improve PTSD symptoms with and without comorbid depression using high and low frequencies applied to left, right, or bilateral sites (Berlim & Van Den Eynde, 2014; Harris & Reece, 2021; Kan et al., 2020; Karsen et al., 2014; Petrosino et al., 2021; Yan et al., 2017; Philip et. al. 2019). The most recent meta-analysis of rTMS in patients with PTSD (Harris & Reece, 2021) demonstrated large improvements in PTSD symptoms (19 studies, 376 subjects, d=1.17, 95% CI 0.89-1.45, p<0.001). A recent retrospective analysis of treatment outcomes in Veterans treated at multiple VA facilities across the country revealed that 70% of the Veterans in this analysis had comorbid PTSD, with the vast majority being treated with 10 Hz rTMS applied to the left DLPFC (Madore et al., 2022). They found significant improvement in depression and PTSD symptoms as measured by the PHQ-9 and PCL-5, respectively. Strikingly, among Veterans with comorbid PTSD, 65.3% experienced clinically meaningful reduction and 46.1% no longer met PTSD criteria (Madore et al., 2022). This cohort study serves as proof of concept for the efficacy of left DLPFC-targeted rTMS in veterans with depression and PTSD.

In trials aiming to alleviate PTSD symptoms, the use of right-sided 5 Hz rTMS has shown promise (Philip et al., 2019). Specifically, a 5 Hz stimulation protocol (which is a distinct and different treatment then intermittent TBS) applied to the right DLPFC was shown to be significantly superior to sham stimulation (Philip et al., 2019). This superiority was demonstrated through improved PTSD symptoms as measured by PCL and CAPS, depression as measured by the inventory of depressive symptomatology-self-report (IDS-SR), as well as social and occupational function. The same authors compared the efficacy of right-sided iTBS to that of 5 Hz rTMS, both targeting the left DLPFC, in a small retrospective study (n=20) of Veterans receiving care at the Providence VA Healthcare System (Philip et al., 2022). Surprisingly the authors found that while both protocols were well tolerated and significantly effective in symptom reduction, iTBS produced inferior outcomes for PTSD (Treatment protocol x Time p=0.011). Intermittent TBS also provided smaller effect sizes than 5Hz rTMS for both PTSD and depression as measured through the PCL-5 and IDS-SR respectively. While this study was targeting right DLPFC and had an overall small sample size, it raises concern for whether an iTBS protocol is suboptimal for patients with depression and concurrent PTSD more generally and re-iterates the importance of determining optimized treatment options for patients with comorbid PTSD and depression.

The VA San Diego Healthcare System (VASDHS) has been providing rTMS to eligible patients with TRD since early 2018. The program primarily utilized 10 Hz rTMS, targeted to the left DLPFC. After the FDA cleared iTBS to be used for TRD, the program switched to primarily offering iTBS targeting the left DLPFC starting in October 2019. Given the well documented efficacy of rTMS for PTSD and the high prevalence of comorbid PTSD and depression in our patient sample, we consistently measured symptoms of both depression and PTSD in our clinic. In this retrospective case-series, we investigated whether there was any difference in efficacy between iTBS and 10 Hz rTMS for treatment of both depression and comorbid PTSD in Veterans. We hypothesized that there would be no difference between stimulation protocols on changes in depression symptoms, thus replicating the initial THREE-D trial (Blumberger et. al, 2018) and further hypothesized that there would be no significant difference in alleviating PTSD symptoms.

## Methods

Data reported following STROBE guidelines.

### Veterans included in analysis

This study was approved as an institutional review board (IRB)-exemption by the VA San Diego Medical Center IRB committee. We conducted a chart review of patients referred to the VA San Diego neuromodulation program who, after consultation, were deemed appropriate for a trial of rTMS (between Feb 2018 through May 2022). Veterans were included for analysis if they received at least 2 weeks of treatment and for whom we had at least 2 measures of their PHQ-9/PCL-5 (with the first acquired within the first week of treatment). Data from 98 Veterans was included in this analysis (see **Table 1** for more details).

**Table 1:**
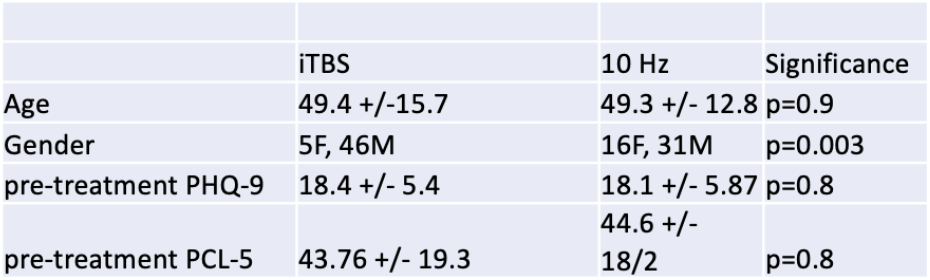
Differences between patients receiving either stimulation protocol. We collected information related to age, gender, pre-treatment PHQ-9 and PCL-5. We only found a significant difference by gender (2-sided Chi-squared test).

All Veterans were initially referred to the VA San Diego neuromodulation program by their primary psychiatrists for an evaluation and potential treatment with either rTMS, electroconvulsive therapy (ECT) or ketamine. Veterans included in this retrospective analysis may thus have been started on rTMS upon initial referral to this program or could have been switched to rTMS after not adequately responding to some other treatment. Eligibility criteria for referral was generally a failure of at least 2 antidepressants in this episode though this criterion was not rigorously upheld in every case. Exclusion criteria for rTMS followed standard guidelines for rTMS: no history of seizures/seizure disorder; no metallic/electrical objects implanted above the head/neck; lack of imminent suicidality and lack of recent substance dependence/misuse (within the last 2 months). Repetitive TMS in our clinic during this period was generally performed 5 days/week, with an initial treatment course defined as 30 treatments.

### Assessments

All data analyzed in this manuscript were gathered as part of clinical care consistent with the clinical program protocol. Veterans were administered a Patient Health Questionnaire-9 (PHQ-9) to monitor symptoms of depression and a Patient Check List-5 (PCL-5) to monitor symptoms of PTSD symptoms prior to the start of rTMS treatment. Scales were then collected weekly after the start of treatment. The PHQ-9 was first developed and validated as a tool for screening for depression in primary care settings, but has been tested and validated in both psychiatric populations more generally (Beard et al., 2016) and as a tool to measure depression-related symptoms at the VA (Katz et al., 2021). The PCL-5, a DSM-V updated version of the PCL, is one of the most widely used self-reported measures of post-traumatic-stress disorder. It has been validated for use in Veterans (Blevins et al., 2015), and is often deployed clinically within the VA system as an easy measure of PTSD severity. We performed a chart review to gather auxiliary data that included age, gender, and formal PTSD diagnosis.

### Statistical Analyses

We focused on three primary questions in this paper:

1. Were there statistically significant and clinically meaningful differences in reductions of either PHQ-9 or the PCL-5 summary score by stimulation type. To analyze this, we implemented a mixed effects 2×2 analysis of variance (ANOVA) model, with time as a within-group repeated measure (pre-stim as T1 and final treatment as T2) and stimulation type as a between-group factor, for both the PHQ-9 and the PCL-5 symptom scales. For T2, we used the symptom score at treatment 30, or else their last reported score if they stopped treatment early (n=98).
2. Would our results vary if restricted to veterans with clinically verified and active PTSD. We repeated analyses in Veterans with both clinically verified PTSD diagnoses (i.e., a listed diagnosis of PTSD in the medical record) and with clinically significant PTSD symptoms (PCL-5 score > 33, prior to starting rTMS treatment, n = 56).
3. We measured whether baseline PTSD symptom scores were a predictor (positive or negative) for antidepressant response using an ANOVA model with change in PHQ-9 as the outcome and PTSD as a fixed factor and a Pearson regression between PCL-5 pre-treatment values and change in PHQ-9 values.

We additionally looked at differences in the Veterans receiving different stimulation types for age, baseline PHQ-9 and PCL-5 scores (using t-tests), and gender (using the Chi-squared test).

### Statistical analyses and reporting

For all ANOVA analyses reported here, we followed the following steps: 1) Normality of score distributions was tested prior to performing further statistical testing using the Shapiro-Wilk test (p>0.05 suggestive of normal distribution). 2) All models were interpreted using a significance level (alpha) of 0.05. 3) For ANOVA models we reported the F statistic (F). Repeated-measures ANOVA used Greenhouse-Geisser correction. For all tests (t-test, regression, Chi-squared test) we report two-sided p values.

## Results

Pre-treatment information that was collected for this manuscript is included in **Table 1**. We first examined whether there was a difference in baseline PHQ-9 or PCL-5 scores between stimulation types. We observed no significant differences in either baseline PHQ-9 scores (t(96) = .25, p = 0.8) or in the PCL-5 scores (t(96) = 0.2 p = 0.8) between the 10 HZ and iTBS groups. There were also not significant differences in age between groups, but there was a significant difference in gender distribution, with 5 females (46 males) in the 10 Hz group and 16 females (31 males) in the iTBS group (p=0.003, Chi-squared test).

Our initial analysis of changes in PHQ-9 scores with treatment showed that both iTBS and 10 Hz resulted in significant and clinically meaningful reductions in depression (**Figure 1A)**. A mixed effects ANOVA was performed to measure whether the change in PHQ-9 scores differed by stimulation type. The model specification included between group factors of stimulation type (10 Hz vs. iTBS) and gender (male vs. female) and a repeated measures factor of time (pre vs. post treatment PHQ-9 score). There was an overall effect of time (F(1,94)=82.5, p<0.001, partial eta-squared = 0.47) and a significant effect by gender (F(1,94)=4.5, p<0.05, partial eta-squared = 0.046). A post-hoc analysis of the gender x time effect revealed that females (n=21) showed a change in PHQ-9 symptoms from 17.1 +/- 1.4 to 8.4 +/- 1.6, while males showed a change from 18.1 +/- 0.65 to 12.7 +/- 0.76. Thus, females in our analysis showed a larger overall reduction in depression symptoms as a consequence of rTMS. There was no time x stimulation type effect (F(1,94) = 0.16, p = 0.69), or time x gender x stimulation type effect (F(1,94)=1, p = 0.32), suggesting that stimulation type did not affect treatment outcome. Post-hoc analyses in this model showed that, after accounting for gender differences, the group receiving iTBS showed a reduction in PHQ-9 scores of 16.7 +/- 1.31 down to 9.4 +/- 1.38 (a 7.3 point reduction), while the group receiving 10Hz treatment showed a reduction in the PHQ-9 of 18.5 +/- 0.86 down to 11.78 +/- 1 (a 6.7 point reduction).

**Figure 1.**
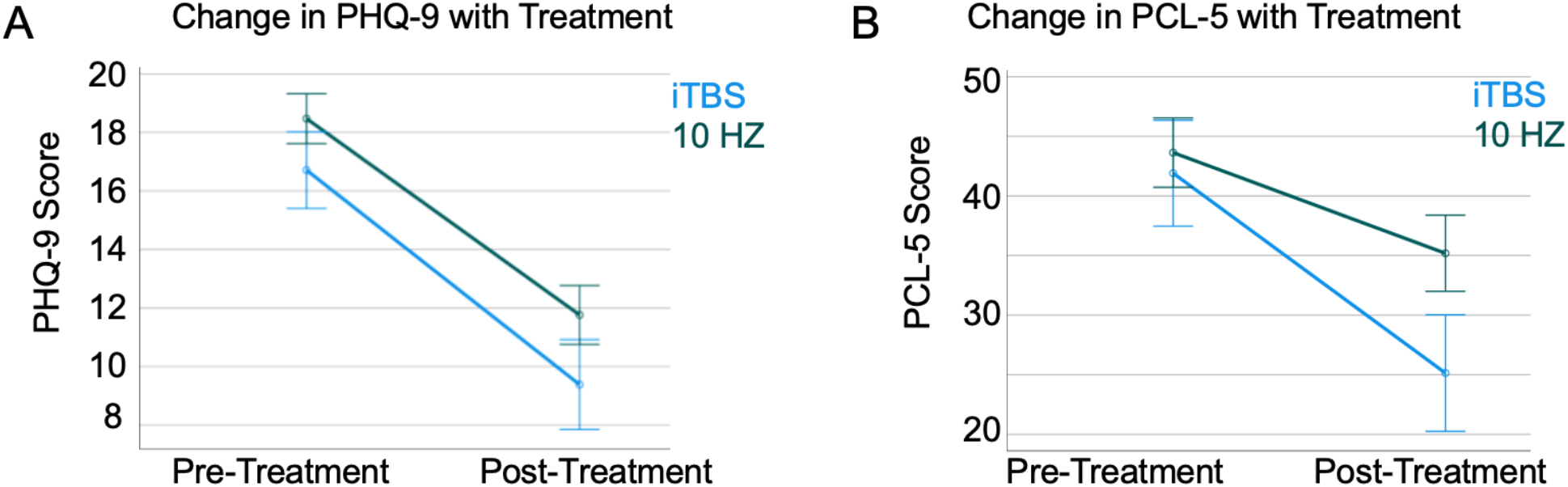
Differences between stimulation protocols in treatment outcomes. We measured change from pre to post-treatment in the PHQ-9 and the PCL-5 separated by stimulation type (data shows mean+/- SEM). We did not observe significant differences in treatment outcomes as a function of stimulation type (P>0.05 for both symptom scales).

We next analyzed changes in PCL-5 scores. We found that both iTBS and 10 Hz stimulation resulted in significant and clinically meaningful reductions in PTSD symptoms as measured using the PCL-5 (**Figure 1B)**. A mixed effects model was specified as above with between group factors of stimulation type and gender and a repeated measures factor of time. This model showed that, for PTSD symptoms, the two types of treatments were similar. There was an overall effect of treatment (F(1,94) = 31.8, p<0.001, partial eta squared = .25). There was not a significant gender x time effect (F(1,94)=1.5, p = 0.2), stimulation type x time effect (F(1,94)=3.46, p = 0.07, partial eta squared = 0.036) or gender x time x stimulation type effect (F(1,94)=0.55, p = 0.46). Follow-up post-hoc analyses showed that the iTBS group showed a reduction from 41.9 +/- 4.4 to 25.1 +/- 4.9 (a difference of 16.8 points) while the 10Hz group showed a reduction from 43.6 +/- 2.9 down to 35.2 +/- 3.2 (a difference of 8.4 points). Thus, we found no evidence that iTBS was inferior to 10Hz treatment in the reduction of PTSD symptoms.

We followed this up by analyzing changes in the PCL-5 with treatment only in Veterans with documented, clinically active PTSD (i.e. diagnosis of PTSD in the chart and a pre-treatment PCL-5 score > 33). In this smaller sample (n=56 total, 31 with iTBS and 25 with 10 Hz treatment, with 12 females and 44 males) we found generally similar effects as reported above. There was similar overall effect of treatment (F(1,52)= 36.5, p<0.001), but no difference by stimulation type (F(1,54)=2.4, p = 0.13) for the change in PCL-5 symptoms. Again, as noted above, post-hoc analysis demonstrated a 20.7 +/- 4.5 point reduction in the iTBS group compared to 12.3 +/- 3.1 point reduction in the 10 Hz group. Thus, we observed no differences between iTBS and standard 10 Hz treatment in the reduction of PTSD symptoms in Veterans, using either the full cohort of Veterans or only those with documented clinically active symptoms of PTSD.

In our final analysis, we measured whether there was an overall relationship between pre-treatment PTSD and antidepressant outcomes, as an earlier study (Yesavage et al. 2018), suggested this. We performed two analyses to measure this. First, using our designation of “clinically active PTSD” noted above, we performed an ANOVA model using change in PHQ-9 as our outcome measure and the presence of clinically active PTSD as a fixed factor (56 Veterans with clinically active PTSD, 42 Veterans without). This model did not show a significant effect of PTSD on the change in PHQ-9 scores with treatment (F (1,95)=0.85, p = 0.4). We performed a follow-up analysis in which we conducted a Pearson’s regression between PCL-5 pre-treatment scores and change in PHQ-9 scores. This model was likewise not significant (r = 0.02, p = 0.9). Thus, there is no evidence in our data that PTSD symptoms, either as a binary (absence or presence) or as a function of symptom severity (correlation analysis), was related to change in depression symptoms.

## Discussion

In this study, we investigated whether there were any differences in iTBS vs. 10 Hz rTMS treatment in overall reduction of depression and PTSD symptoms among Veterans. We found no clinically meaningful differences in the reduction in either depression or PTSD symptoms based on the type of stimulation. Our findings are consistent with the large THREE-D trial (Blumberger et. al., 2018), though extend this finding to a Veteran population, and to PTSD symptoms as well as depression symptoms. Our findings are also generally consistent with other recent studies from the VA showing that rTMS can improve symptoms of PTSD, though prior reports focused mostly on standard 10Hz stimulation protocols (Madore et al., 2022; Wilkes et al., 2020). Our findings, that rTMS can improve PTSD symptoms in Veterans are also consistent with those from an RCT determining the efficacy of active iTBS targeted to right-DLPFC on PTSD symptoms, as the authors found significant improvement in both PTSD and depression from the active arm of the trial at one month follow up (Philip et al., 2019). This trial’s patient population mirrored ours as it included Veterans with PTSD, most of whom (90%) also had comorbid depression.

Our outcomes differ from those observed by Philip et al. (2022), who found that different iTBS targeted to right-DLPFC was less effective than 5Hz stimulation in reducing PTSD symptoms. Their study was distinct from our in several notable ways. First and most importantly, their study compared different stimulation of 5Hz vs. iTBS targeted to the right hemisphere, whereas we were comparing 10Hz and iTBS targeted to the left hemisphere. While there is no clearly identified treatment or optimal stimulation frequency for PTSD (as reviewed thoroughly here: Petrosino et al., 2021; Kozel et. al. 2019), there is evidence that right sided stimulation may be more effective than left (Boggio et al., 2010). Thus, the differences observed in the Philip et. al. 2022 study may be related to differences between right vs. left-sided stimulation. Also, both our and their studies has small samples (the above study had a total of 20 subjects and even our study only had 56 Veterans with clinically verifiable PTSD symptoms), and both suffer from non-randomization and non-blinded protocols. Thus, further research using appropriate controls/blinding and randomization is needed. However, our study at least suggests that such research is warranted.

Contrary to the findings of Yesavage et al. (2018), we did not find that PTSD symptoms (as defined by both formally diagnosed and pre-treatment PCL-5 values >33) were associated with less improvement in depression scores either categorically (as defined by both formally diagnosed and pre-treatment PCL-5 values >33) or as a function of symptom scores. Rather our study participants and results are consistent with those from Hernandez et al., (2020) and Wilkes et al., (2020). In both studies, Veterans with depression and comorbid PTSD treated with rTMS experienced significant improvements in both disorders, with similar improvements in their depression symptoms compared to Veterans without PTSD.

Intermittent TBS shows several clear advantages over 10Hz treatments generally. First, accelerated TMS protocols (in which multiple treatments are delivered on the same day) were recently FDA-cleared for the treatment of depression (Cole et al., 2020). Accelerated protocols rely on short iTBS treatments to adequately space out repeated treatments. Our results support the idea that accelerated protocols may be useful in PTSD as well and warrant further research in this area. In addition, psychotherapy is fundamental to the treatment of PTSD, and recently various trials have been published combining rTMS with various forms of psychotherapy. For instance, an RCT has shown that 10 Hz rTMS in conjunction with cognitive processing therapy (CPT) yields a significantly greater improvement than sham stimulation and CPT (Kozel et al., 2018). It is possible that iTBS, due to shorter stimulation periods, will allow for improved or novel treatment paradigms in which TMS can be combined with various forms of psychotherapy.

There are limitations to our study that need to be factored into the interpretation of the findings. First, this is a retrospective and non-randomized design and is associated with the usual challenges of interpreting this data. Due to the non-random allocation, the two groups of subjects may possess different qualities or attributes. Among these differences, patient sex warrants further discussion. Analysis of over 5,000 patients treated with TMS for depression found female participants more likely to respond to treatment and enter remission (Sackeim et al., 2020). Several possible biological explanations stemming from differences in patient sex have since been described (Hanlon & McCalley, 2022). We reported unbalanced gender distribution among the two groups, with the 10 Hz group having more women overall and as a higher proportion of participants. We did indeed show an effect of gender and included this as a factor in our models, but these differences would be better accounted for by proper randomization. The groups for the most part underwent treatment across non overlapping periods and importantly, the COVID-19 pandemic overlapped with the treatment delivered during the iTBS period. The psychiatric burden of the pandemic could have confounded the baseline of the group receiving iTBS. The treatments used different devices: 10 Hz treatments were delivered using a MagStim device, while iTBS treatments were delivered using a MagVenture device. Other limitations include a medium sample-size and thus a limitation in the effect size difference we were powered to observe. Finally in this study we were not specifically treating Veterans with PTSD, rather Veterans who initially presented for treatment of depression, and effects may differ in Veterans who are primarily presenting for treatment for PTSD symptoms. However, studies show that the majority of Veterans with resistant depression have comorbid PTSD, and thus our results are likely relevant to many Veterans receiving treatment.

Considering these limitations, there are several positive aspects to our study. First, despite all of these complexities, it is important to note that we did replicate the overall effects observed previously in the large, randomized/blinded THREE-D trial (Blumberger et al., 2018). This allows us to have greater confidence in our analysis of PTSD outcomes. Second, while we did not randomize Veterans to the two treatments, there was a “pseudo-randomization” built in to how the treatments were offered (for 2 years, only 10Hz treatments were offered; and after we switched iTBS became the default treatment of choice in our clinic). This somewhat mitigates the lack of randomization, in that it was less likely that Veteran characteristics or some other aspect related to depression severity itself influenced which treatment was offered. Finally, there is no evidence to date that outcomes differ based on manufacturer type, given similar magnet geometries. Indeed, FDA-clearance for these different devices are predicated on them being overall similar in their ability to provide brain stimulation.

## Data Availability

All data produced in the present study are available upon reasonable request to the authors

